# Deep Learning-based Detection for COVID-19 from Chest CT using Weak Label

**DOI:** 10.1101/2020.03.12.20027185

**Authors:** Chuansheng Zheng, Xianbo Deng, Qiang Fu, Qiang Zhou, Jiapei Feng, Hui Ma, Wenyu Liu, Xinggang Wang

## Abstract

Accurate and rapid diagnosis of COVID-19 suspected cases plays a crucial role in timely quarantine and medical treatment. Developing a deep learning-based model for automatic COVID-19 detection on chest CT is helpful to counter the outbreak of SARS-CoV-2. A weakly-supervised deep learning-based software system was developed using 3D CT volumes to detect COVID-19. For each patient, the lung region was segmented using a pre-trained UNet; then the segmented 3D lung region was fed into a 3D deep neural network to predict the probability of COVID-19 infectious. 499 CT volumes collected from Dec. 13, 2019, to Jan. 23, 2020, were used for training and 131 CT volumes collected from Jan 24, 2020, to Feb 6, 2020, were used for testing. The deep learning algorithm obtained 0.959 ROC AUC and 0.976 PR AUC. There was an operating point with 0.907 sensitivity and 0.911 specificity in the ROC curve. When using a probability threshold of 0.5 to classify COVID-positive and COVID-negative, the algorithm obtained an accuracy of 0.901, a positive predictive value of 0.840 and a very high negative predictive value of 0.982. The algorithm took only 1.93 seconds to process a single patient’s CT volume using a dedicated GPU. Our weakly-supervised deep learning model can accurately predict the COVID-19 infectious probability in chest CT volumes without the need for annotating the lesions for training. The easily-trained and highperformance deep learning algorithm provides a fast way to identify COVID-19 patients, which is beneficial to control the outbreak of SARS-CoV-2. The developed deep learning software is available at https://github.com/sydney0zq/covid-19-detection.

## 1 Introduction

Since Dec 2019, a large and increasing outbreak of a novel coronavirus was first emerging in Wuhan, Hubei province of China [1, 2], which can cause acute respiratory illness and even fatal acute respiratory distress syndrome (ARDS) [3]. The new coronavirus was named as SARS-CoV-2 by International Committee on Taxonomy of Viruses (ICTV) [4] and the infectious diseases infected by this coronavirus was named as Coronavirus Disease 2019 (COVID-19) by World Health Organization (WHO) [5]. The new coronavirus has been confirmed of human-to-human transmission [6, 7], and due to the massive transportation and large population mobility before the Chinese Spring Festival, this new coronavirus has spread fast to other areas in China with considerable morbidity and mortality. According to the data from the National Health Commission of the People’s Republic of China [8], update till 24 o’clock of Mar 6, 2020, China has reported 80651 identified cases with SARS-CoV-2, including 3070 death cases; 83.9% (67666/80651) of the identified cases came from Hubei province and identified cases in Wuhan accounted about 73.7% (49871/67666) of the data in Hubei province. Moreover, several exported cases outside China have been reported in more than 20 countries, such as Iran, Japan, South Korea, USA, Singapore, Germany, Vietnam, Thailand, and some other countries, and some person-to-person spread of this new virus has also been detected. With the risk of further spread of SARS-CoV-2, it has been declared to be a Public Health Emergency of International Concern (PHEIC) by WHO on 30 January 2020 [4], which poses a great threat to the international human health.

Even though real-time reverse transcriptase polymerase chain reaction (RT-PCR) has been considered as the gold standard for SARS-CoV-2 diagnosis, the very limited supply and strict requirements for laboratory environment would greatly delay accurate diagnosis of suspected patients, which has posed unprecedented challenges to prevent the spread of the infection, particularly at the center of the epidemic area. In contrast with it, chest computed tomography (CT) is a faster and easier method for clinical diagnosis of COVID-19 by combining the patient’s clinical symptoms and signs with their recent close contact, travel history, and laboratory findings, which can make it possible for quick diagnosis as early as possible in the clinical practice. It is also effectively helpful to isolate infected patients timely and control the epidemic, especially within the range of Wuhan, Hubei province. In a word, chest CT is a key component of the diagnostic procedure for suspected patients and its CT manifestations have been emphasized in several recent reports [1, 9, 10, 11, 12].

In a word, accurate and rapid diagnosis of COVID-19 suspected cases at the very early stage plays a crucial role in timely quarantine and medical treatment, which is also of great importance for patients’ prognosis, the control of this epidemic, and the public health security. But currently, a large number of suspected patients need to undergo the chest CT scanning in Hubei province, which have caused a tremendous burden to professional medical staffs, and their severe shortage is also a major challenge in the current situation; moreover, radiologists’ visual fatigue would heighten the potential risks of missed diagnosis for some small lesions.

Deep learning, as the core technology of the rising artificial intelligence (AI) in recent years, has been reported with significantly diagnostic accuracy in medical imaging for automatic detection of lung diseases [13, 14, 15]. It surpassed human-level performance on the ImageNet image classification task with one million images for training in 2015 [16], showed dermatologist-level performance on classifying skin lesions in 2017 [17] and obtained very impressive results for lung cancer screening in 2019 [13]. However, most deep learning based methods for disease diagnosis requires to annotate the lesions, especially for disease detection in CT volumes. In the current, annotating lesions of COVID-19 costs a huge amount of efforts for radiologists, which is not acceptable when COVID-19 is spreading fastly and there are great shortages for radiologists. Thus, performing COVID-19 detection in a weakly-supervised manner is of great importance. One of the simplest labels for COVID-19 detection is the patient-level, i.e., indicating the patient is COVID-19 positive or negative. Therefore, aim of current study was to investigate the potential of a deep learning-based model for automatic COVID-19 detection on chest CT volumes using the weak patient-level label, for the sake of rapid diagnosis of COVID-19 at this critical situation to help to counter this outbreak, especially within Wuhan, Hubei province, China.

## 2 Material and methods

### Patients

This retrospective study was approved by Huazhong University of Science and Technology ethics committee, patient consent was waived due to the retrospective nature of this study.

Between Dec. 13, 2019 to Feb. 6, 2020, we searched unenhanced chest CT scans of patients with suspected COVID-19 from the picture archiving and communication system (PACS) of radiology department (Union Hospital, Tongji Medical College, Huazhong University of Science and Technology). Finally, 540 patients (mean age, 42.5 ± 16.1 years; range, 3-81 years, male 226, female 314) were enrolled into this study, including 313 patients (mean age, 50.7 ± 14.7 years; range, 8-81 years; male 138, female 175) with clinical diagnosed COVID-19 (COVID-positive group) and 229 patients (mean age, 31.2±10.0 years; range, 3-69 years; male 88, female 141) without COVID-19 (COVID-negative group). There was no significant difference in sex between the two groups (*χ*^2^=1.744; P=0.187), age in COVID-positive group significantly higher than that of COVID-negative group (t=17.09; P<0.001). The main clinical symptoms for these patients were fever, cough, fatigue, and diarrhea. Of all the patients, two were included by both groups due to the first and second follow-up CT scans. The first case (female, year 66) was diagnosed as COVID-19 negative on Jan 24, 2020, then changed into COVID-positive on Feb 6, 2020; the second case (female, year 23) was diagnosed as COVID-19 positive on Jan 24, 2020, then changed into COVID-negative on Feb 3, 2020. All the CT volumes scanned on and before Jan 23, 2020, were assigned for deep learning training, and all the CT volumes scanned after Jan 23, 2020, were assigned for deep learning testing.

### Image Acquisition

The CT scanning of all the enrolled patients was performed on a gemstone CT scanner (GE Discovery CT750HD; GE Healthcare, Milwaukee, WI), and were positioned in a headfirst supine position, with their bilateral arms raised and placed beside bilateral ears. All the patients underwent CT scans during the end-inspiration without the administration of contrast material. Related parameters for chest CT scanning were listed as follows: field of view (FOV), 36 cm; tube voltage, 100 kV; tube current, 350 mA; noise index, 13; helical mode; section thickness, 5 mm; slice interval, 5 mm; pitch, 1.375; collimation 64 × 0.625 mm; gantry rotation speed, 0.7 s; matrix, 512 × 512; the reconstruction slice thickness 1 mm with an interval of 0.8 mm; scan rage from apex to lung base; the mediastinal window: window width of 200 HU with a window level of 35 HU, and the lung window: window width of 1500 HU with a window level of -700 HU.

### Ground-truth Label

In the latest diagnosis and treatment protocols of pneumonia caused by a novel coronavirus (trial version 5) [18] which was released by National Health Commission of the People’s Republic of China on Feb 4, 2020, suspected cases with characteristic radiological manifestations of COVID-19 has been regarded as current standard for clinical diagnostic cases in severely affected areas only in Hubei Province, indicating that chest CT is fundamental for COVID-19 identification of clinically diagnosed cases.

Typical CT findings for COVID-19 are also listed: multifocal small patchy shadowing and interstitial abnormalities in the early stage, especially for the peripheral area of the bilateral lungs. In the progressive period, the lesions could increase in range and in number; it could develop into multiple ground glass opacity (GGO) with further infiltration into the bilateral lungs. In severe cases, pulmonary diffuse consolidation may occur and pleural effusion is rarely shown.

The combination of epidemiologic features (travel or contact history), clinical signs and symptoms, chest CT, laboratory findings and real-time RT-PCR (if available) for SARS-CoV-2 nucleic acid testing is used for the final identification of COVID-19. The medical CT reports were acquired via the electronic medical record of Union Hospital, Tongji Medical College, Huazhong University of Science and Technology. According to the CT reports, if a CT scan was COVID-positive, its ground-truth label was 1; otherwise, the label was 0.

### The Proposed DeCoVNet

We proposed a 3D deep convolutional neural Network to Detect COVID-19 (DeCoVNet) from CT volumes. As shown in Fig. 1, DeCoVNet took a CT volume and its 3D lung mask as input. The 3D lung mask was generated by a pre-trained UNet [19]. DeCoVNet was divided into three stages for a clear illustration in Table. 1. The first stage was the network stem, which consisted of a vanilla 3D convolution with a kernel size of 5 × 7 × 7, a batchnorm layer and a pooling layer. The second stage was composed of two 3D residual blocks (ResBlocks). In each ResBlock, a 3D feature map was passed into both a 3D convolution with a batchnorm layer and a shortcut connection containing a 3D convolution that was omitted in Fig. 1 for dimension alignment. The resulted feature maps were added in an element-wise manner. The third stage was a progressive classifier (ProClf), which mainly contained three 3D convolution layers and a fully-connected (FC) layer with the softmax activation function. ProClf progressively abstracts the information in the CT volumes by 3D max-pooling and finally directly output the probabilities of being COVID-positive and COVID-negative.

**Table 1:** Detailed structure of the proposed DeCovNet. The number after the symbol “@”, e.g., 5 × 7 × 7, denotes the kernel size of the convolution layer or the residual block. “&” means that there are two types of kernel size in the residual block. “T” denotes the length of the input CT volume. The number in “Output size” is in the order of “channel, length, height, width”. The input size is 2 × *T* × 192 × 288.

| Stages | Layers | Output size |
| --- | --- | --- |
| Stem | Conv3d(2, 16)@$5 \times 7 \times 7$+BN+ReLU | $16 \times T \times 48 \times 72$ |
| ResBlocks | ResBlock(16, 64)@$3 \times 1 \times 1$&$1 \times 3 \times 3$ | $64 \times T \times 48 \times 72$ |
| | MaxPool3d | $64 \times T/2 \times 48 \times 72$ |
| | ResBlock(64, 128)@$3 \times 1 \times 1$&$1 \times 3 \times 3$ | $128 \times T/2 \times 24 \times 36$ |
| Progressive classifier | AdaptiveMaxPool3d | $128 \times 16 \times 24 \times 36$ |
| | Conv3d(128, 64)@$3 \times 3 \times 3$+ReLU | $64 \times 16 \times 24 \times 36$ |
| | AdaptiveMaxPool3d | $64 \times 4 \times 12 \times 18$ |
| | Conv3d(64, 32) @$3 \times 3 \times 3$+ReLU | $32 \times 4 \times 12 \times 18$ |
| | Dropout3d(p=0.5) | $32 \times 4 \times 12 \times 18$ |
| | Conv3d(32, 32)@$3 \times 3 \times 3$+ReLU | $32 \times 4 \times 12 \times 18$ |
| | AdaptiveMaxPool3d | $32 \times 1 \times 1 \times 1$ |
|  | FullyConnected(32, 2) | 2 |

**Figure 1:**
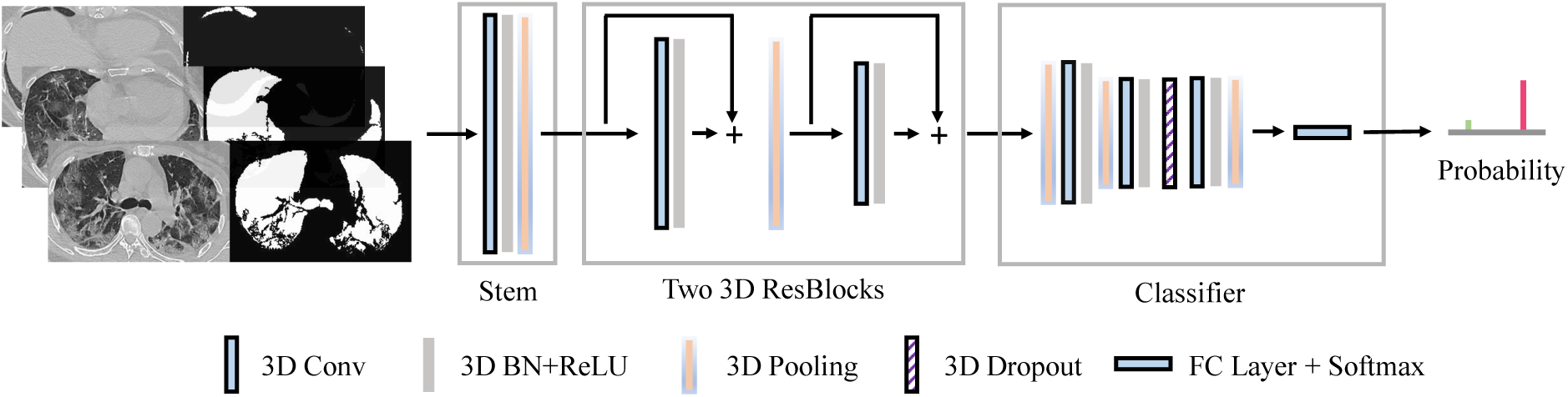
Architecture of the proposed DeCoVNet. The network took a CT volume with its 3D lung mask as input and directly output the probabilities of COVID-positive and COVID-negative.

The 3D lung mask of an input chest CT volume helped to reduce background information and better detect COVID-19. Detecting the 3D lung mask was a well-studied issue. In this study, we trained a simple 2D UNet using the CT images in our training set. To obtain the ground-truth lung masks, we segmented the lung regions using an unsupervised learning method [20], removed the failure cases manually, and the rest segmentation results were taken as ground-truth masks. The 3D lung mask of each CT volume was obtained by testing the trained 2D UNet frame-by-frame without using any temporal information. The overall training and testing procedures of UNet and DeCoVNet for COVID-19 detection were illustrated in Fig. 2.

**Figure 2:**
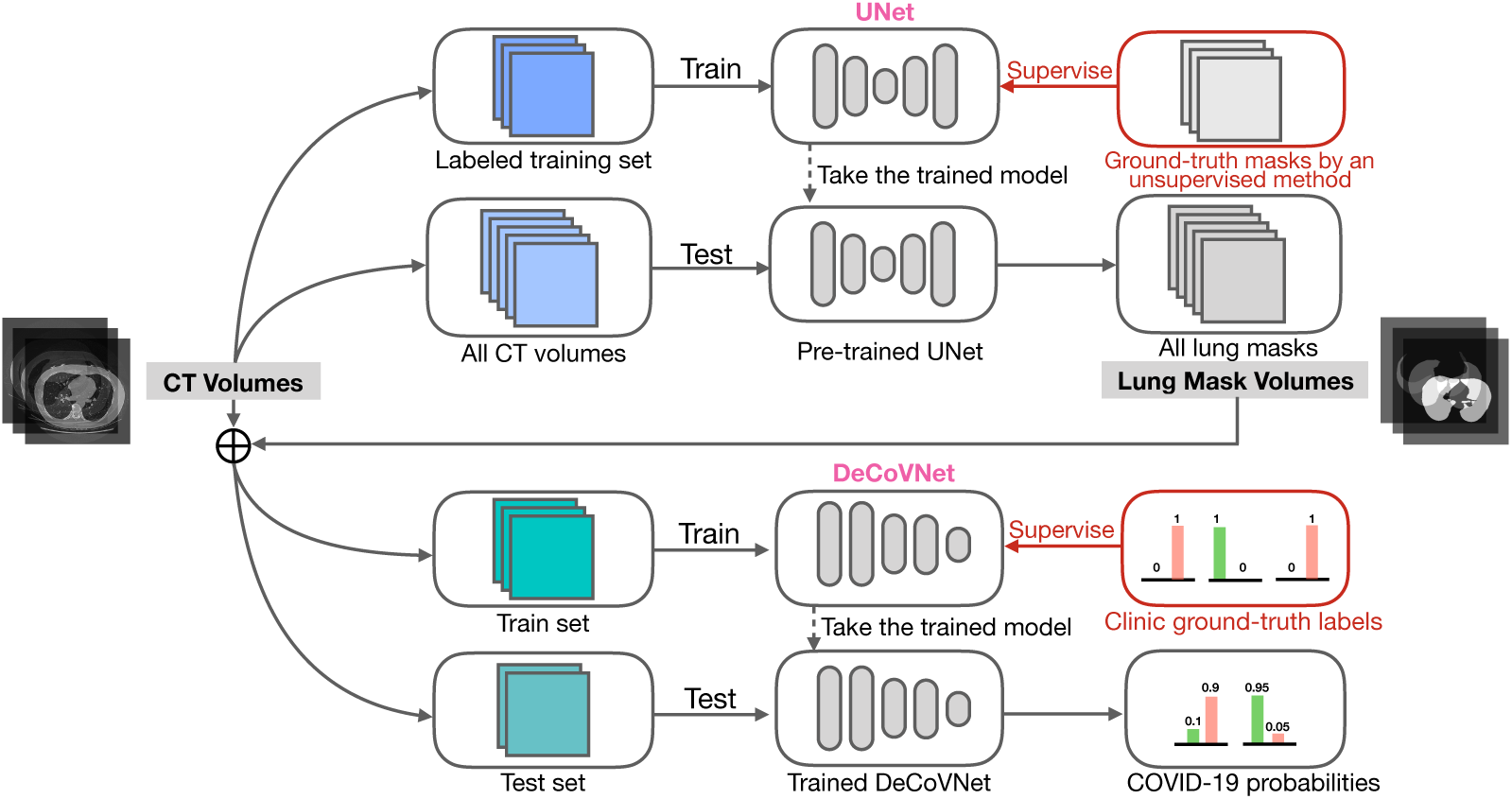
Training and testing procedures. A UNet for lung region segmentation was first trained on the labeled training set using the ground-truth lung masks generated by an unsupervised learning method. Then, all CT volumes were tested by the pre-trained UNet to obtain all lung masks. Each CT volume was concatenated with its lung mask volume as the input of DeCoVNet. DeCoVNet was trained under the supervision of clinical ground-truth labels (COVID-positive and COVID-negative). Lastly, the trained DeCoVNet made predictions on the testing set.

### Data Preprocessing and Data Augmentation

#### Preprocessing of 2D UNet

All the CT volumes were preprocessed in a unified manner before training the 2D UNet for lung segmentation. First, the unit of measurement was converted to the Hounsfield Unit (HU) and the value was linearly normalized from 16-bit to 8-bit (i.e., 0-255) after determining the threshold of a HU window (e.g., -1 200-600 HU). After that, all the CT volumes were resampled into a same spatial resolution (e.g., 368 × 368), by which the CT volumes could be aligned without the influence of the cylindrical scanning bounds of CT scanners. This step was applied to the obtained ground-truth lung masks as well.

#### Preprocessing of DeCoVNet

For each CT volume, the lung masks produced by the trained UNet formed a mask volume, then the CT volume was concatenated with the mask volume to obtain a CT-Mask volume. Finally, the CT-Mask volume was resampled into a fixed spatial resolution (e.g., 224 × 336) without changing the number of slices for DeCoVNet training and testing. The number of slices in the whole dataset was 141±16 ranging from 73 to 250.

#### Data Augmentation

To avoid the overfitting problem since the number of training CT volumes was limited, online data augmentation strategies were applied including random affine transformation and color jittering. The affine transformation was composed of rotation (0°±10°), horizontal and vertical translations (0% ±10%), scaling (0%±20%) and shearing in the width dimension (0°±10°). The color jittering adjusted brightness (0%± 50%) and contrast (0%±30%). For each training sample, the parameters were randomly generated and the augmentation was identically applied for each slice in the sampled CT volume.

#### Training and Testing Procedures

The DeCoVNet software was developed based on the PyTorch framework [21]. Our proposed DeCoVNet was trained in an end-to-end manner, which meant that the CT volumes were provided as input and only the final output was supervised without any manual intervention. The network was trained for 100 epochs using Adam optimizer [22] with a constant learning rate of 1e-5. Because the length of CT volume of each patient was not fixed, the batch size was set to 1. The binary cross-entropy loss function was used to calculate the loss between predictions and ground-truth labels.

During the procedure of testing, data augmentation strategies were not applied. The trained DeCoVNet took the preprocessed CT-Mask volume of each patient and output the COVID-positive probability as well as COVID-negative probability. Then the predicted probabilities of all patients and their corresponding ground-truth labels were collected for statistical analysis.

The cohort for studying the COVID-19 detection contained 630 CT scans collected from Dec 13, 2019 to Feb 6, 2020. To simulate the process of applying the proposed DeCoVNet for clinical computeraided diagnosis (i.e., prospective clinical trials), we used the 499 CT scans collected from Dec 13, 2019 to Jan 23, 2020 for training and used the rest 131 CT volumes collected from Jan 24, 2020 to Feb. 06, 2020 for testing. Of the training volumes, 15% were randomly selected for hyperparameter tuning during the training stage.

#### Statistical Analysis

COVID-19 detection results were reported and analyzed using receiver operating characteristic (ROC) and precision-recall (PR) curves. The area under the ROC curve (ROC AUC) and the area under the precision-recall curve (PR AUC) were calculated. Besides, multiple operating points were chosen on the ROC curve, e.g., the points with approximately 0.95 sensitivity (high sensitivity point) and with approximately 0.95 specificity (high specificity point). ROC AUC, PR AUC, and some key operating points were used to assess the deep learning algorithm.

#### Patient and Public Involvement

This was a retrospective case series study and no patients were involved in the study design, setting the research questions, or the outcome measures directly. No patients were asked to advise on interpretation or writing up of results.

## 3 Experimental results

The software for COVID-19 detection with the pre-trained model as well as the results was available at https://github.com/sydney0zq/covid-19-detection, which will be made publicly available on the publication of this paper. Training DeCoVNet on the training set which consisted of 499 CT volumes took about 20 hours (11 hours for UNet and 9 hours for DeCoVNet) and testing a CT volume costed an average of 1.93 seconds (1.80 seconds for UNet and 0.13 seconds for DeCoVNet) on an NVIDIA Titan Xp GPU.

For every testing CT scan, we used the trained DeCoVNet to predict its probability of COVID-19. By comparing with their binary ground-truth labels, we plotted ROC and PR curves as shown in Fig. 3 and Fig. 4 respectively. In the ROC, we obtained a ROC AUC value of 0.959. When true positive rate (TPR, i.e., sensitivity) was approximately 0.95, our model obtained a true negative rate (TNR, i.e., specificity) of 0.786; when TNR was approximately 0.95, our model obtained a TPR of 0.880; there was another operating showed that our algorithm obtained both TPR and FPR larger than 0.9, i.e., sensitivity=0.907 and specificity=0.911. On the PR curve, our model obtained a PR AUC of 0.975.

**Figure 3:**
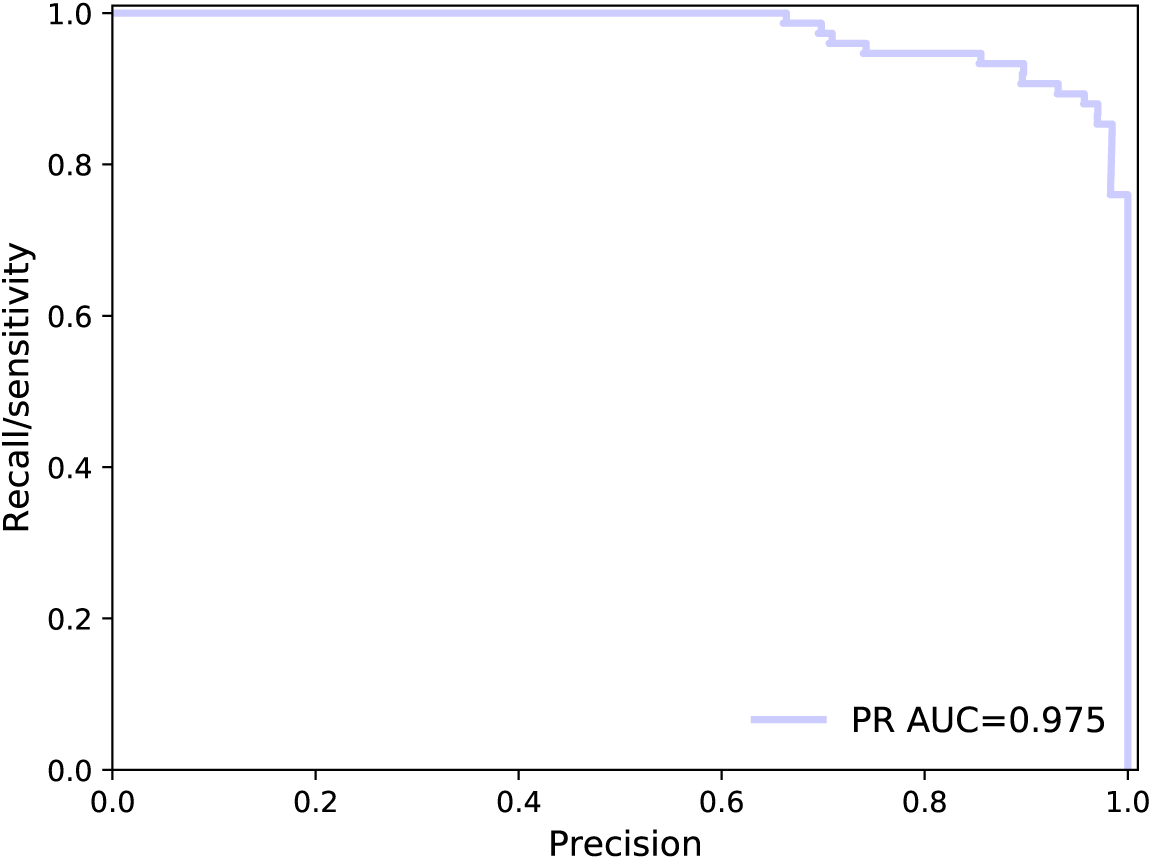
COVID-19 detection results evaluated using the receiver operating characteristic curve.

**Figure 4:**
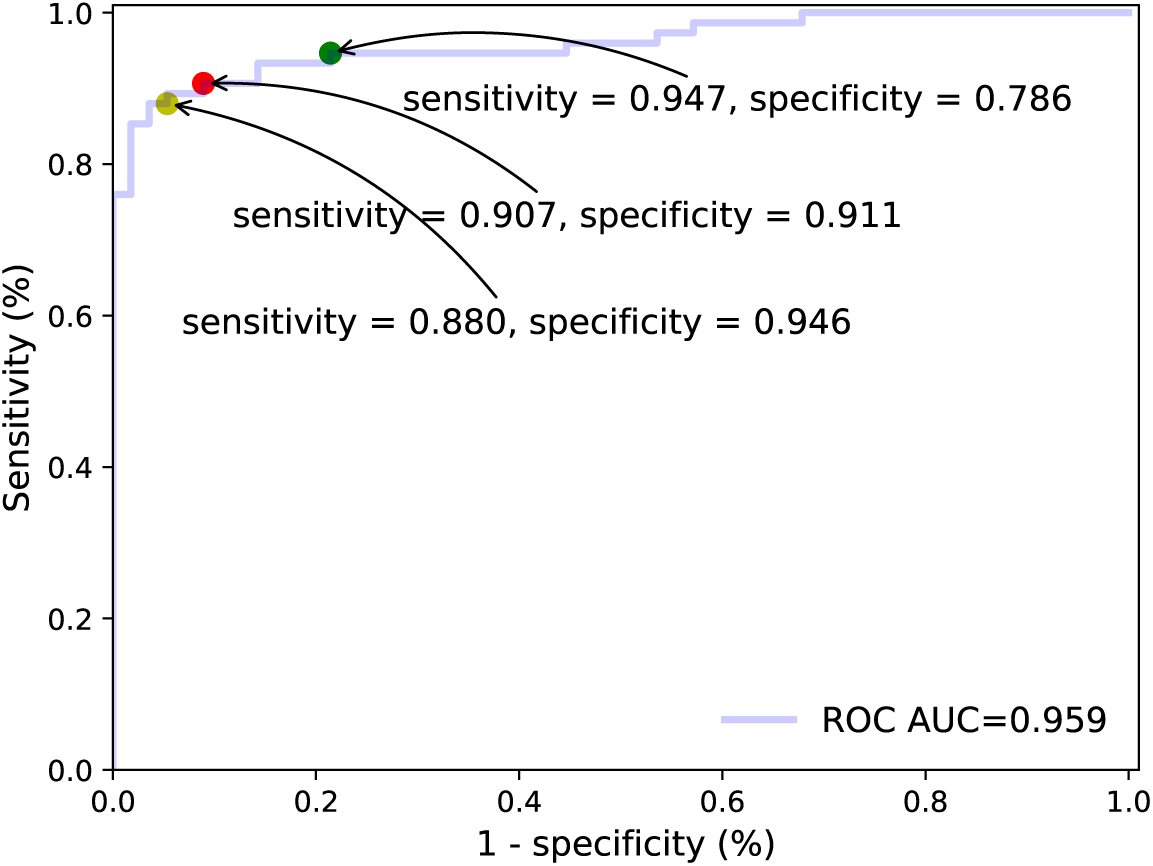
COVID-19 detection results evaluated using the precision-recall curve.

When using the threshold of 0.5 to make COVID-19 detection prediction (i.e., if the probability of COVID-19 was larger than 0.5, the patient was classified as COVID-positive, and vice versa), the algorithm obtained an accuracy of 0.901 with a positive predictive value (PPV) of 0.840 and a negative predictive value (NPV) of 0.982. By varying the probability threshold, we obtained a series of COVID-19 detection accuracy, PPV and NPV in Table 2. Our data showed that the COVID-19 prediction accuracy obtained by the DeCoVNet algorithm was higher than 0.9 when the threshold ranged from 0.2 to 0.5. At the threshold setting of 0.5, there were 12 false positive predictions in total and only one false positive prediction by the algorithm in our study, indicating that the algorithm to have a very high negative predictive value.

**Table 2:** COVID-19 detection statistics by varying the probability thresholds. (PPV: positive prediction value. NPV: negative prediction value.)

| Threshold | Accuracy | PPV | NPV |
| --- | --- | --- | --- |
| 0.1 | 0.893 | 0.933 | 0.839 |
| 0.2 | 0.908 | 0.880 | 0.946 |
| 0.3 | 0.908 | 0.867 | 0.964 |
| 0.4 | 0.901 | 0.840 | 0.982 |
| 0.5 | 0.901 | 0.840 | 0.982 |
| 0.6 | 0.878 | 0.800 | 0.982 |
| 0.7 | 0.870 | 0.787 | 0.982 |
| 0.8 | 0.855 | 0.760 | 0.982 |
| 0.9 | 0.847 | 0.733 | 1.000 |

The accurate predictions (a true positive and a true negative) were presented in Fig. 5 (A-B), and erroneous predictions in Fig. 5 (C-F). In images corresponding to the true positive and the false negative, the lesions of COVID-19 were annotated by red arrows. As shown in Fig. 5 (C, D, E), the false negative predictions were made by the algorithm, and Fig. 5 (F) showed the only false positive prediction, in which the respiratory artifact had been mistaken as a COVID-19 lesion by the DeCoVNet algorithm.

**Figure 5:**
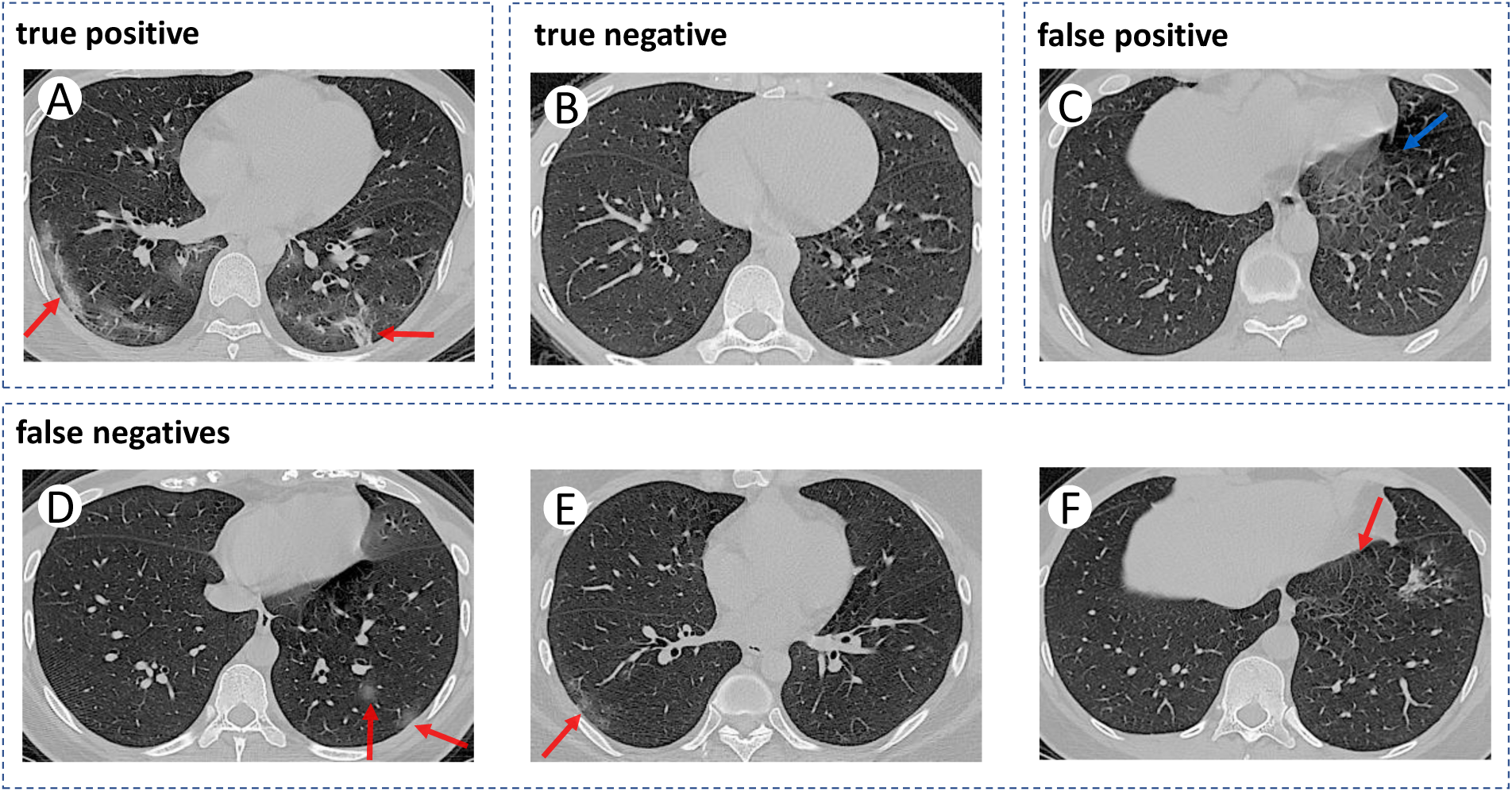
Some accurate and erroneous predictions of the proposed DeCoVNet.

## 4 Discussion

To our knowledge, this is the first study to perform weakly-supervised computer-aided COVID-19 detection with a large number of CT volumes from the frontline hospital at the present epidemic period. By designing an effective weakly-supervised deep learning-based algorithm and training it on CT volumes collected before Jan 23, 2020 with only patient-level labels, the testing results on 131 CT scans collected from Jan 24, 2020, to Feb 6, 2020, were very impressive, e.g., the PR AUC value was 0.975. On the ROC curve, the algorithm obtained sensitivity and specificity values larger than 0.9, which were both clinically applicable.

The motivation of this study was to utilize AI to alleviate the problem of shortage of professional interpretations for CT images when the epidemic is still fast spreading. Though there were many effective applications of medical AI in previous studies [13, 23], developing AI for automatic COVID-19 detection was still a challenging task. Firstly, in the current emergency situation, the number of enrolled patients is relatively smaller compared with that used in previous studies [13, 23]; and patients enrolled in our study were clinically diagnosed cases with COVID-19, because the majority of them did not undergo the nucleic acid testing due to the sudden outbreak and limited medical resource in such a short time period. Secondly, the lesions of COVID-19 in CT volumes were not labeled by radiologists and only patient-level labels (i.e., COVID-positive or COVID-negative) were utilized for training the AI algorithm in our study. Thirdly, some small infected areas of COVID-19 have the potential to be missed even by professional radiologists, and whether it is feasible to be detected by deep learning-based 3D DCNN model remains unclear. We hypothesized to solve these problems by proposing a delicate 3D DCNN, i.e., DeCoVNet. It solved the first problem by applying extensive data augmentation on training CT volumes to obtain more training examples. The second problem was solved by regarding the COVID-19 detection problem as a weakly-supervised learning problem [24], i.e., detecting COVID-19 without annotating the regions of COVID-19 lesions. In the designed DeCoVNet, we used the spatially global pooling layer and the temporally global pooling layer to technically handle the weakly-supervised COVID-19 detection problem. The third problem was addressed by taking the advantages of deep learning and utilizing a pre-trained UNet for providing the lung masks to guide the learning of DeCoVNet.

The deep learning-based COVID-19 diagnostic algorithm used in our study is effective compared to recent deep learning-based computer-aided diagnosis methods. On the task of predicting the risk of lung cancer [13], the deep learning model was trained on 42290 CT cases from 14851 patients and obtained 0.944 ROC AUC. On the task of critical findings from head CT [23], the deep learning model was trained on 310055 head CT scans and obtained ROC AUC of 0.920. In our study, only 499 scans were used for training, but the obtained ROC AUC was 0.959. By comparing the data between them, it was able to find that the task of COVID-19 detection may be easier and the proposed deep learning algorithm was very powerful. As for the erroneous 12 false negative predictions in our results, the most possible explanations after we rechecked the original CT images were listed as follows: those lesions were slightly increased in CT densities, and images of those ground-glass opacities were very faint without consolidation.

Our study provided a typical and successful solution for developing medical AI for emerging diseases, such as COVID-19. While we were developing this AI, doctors in Wuhan were still extremely busy with treating a huge number of COVID-19 patients and it may be impossible for them to annotate the lesions in CT volumes in the current austere fight against this epidemic. Thanks to the weakly-supervised algorithm in this study, locations of pulmonary lesions in CT volumes are not necessary to be annotated, and radiologists’ annotating efforts can be minimized, i.e., only providing patient-level labels. Therefore, developing a helpful AI tool swiftly has become possible and available in the clinical application. In the future, the burden of AI experts could be lifted significantly by automatic machine learning (AutoML) [25].

### Limitations of this study

There are still several limitations in this study. First, network design and training may be further improved. For example, the UNet model for lung segmentation did not utilize temporal information and it was trained using imperfect ground-truth masks, which could be improved by using 3D segmentation networks and adopting precise ground-truth annotated by experts. Second, the data used in this study came from a single hospital and cross-center validations were not performed. Third, when diagnosing COVID-19, the algorithm worked in a black-box manner, since the algorithm was based on deep learning and its explainability was still at an early stage. Related work of all limitations mentioned above will be addressed in our further studies.

## 5 Conclusion

In conclusion, without the need for annotating the COVID-19 lesions in CT volumes for training, our weakly-supervised deep learning algorithm obtained strong COVID-19 detection performance. Therefore, our algorithm has great potential to be applied in clinical application for accurate and rapid COVID-19 diagnosis, which is of great help for the frontline medical staff and is also vital to control this epidemic worldwide.

## Data Availability

No additional data are available.

## Contributors

CZ, XD, QF and QZ contributed equally to this study and are considered as joint first authors. XW, CZ, XD, QZ, QF and LW conceived and designed the study. QZ, XW and JF developed the algorithms with the help of clinical input from CZ, XD and QF. CZ, XD and HM collected, anonymized, and prepared the data from Department of Radiology, Union Hospital, Tongji Medical College, Huazhong University of Science and Technology, China. XD and QF contributed to the protocol of the study. XW and QF did the statistical analysis. XW, QF and QZ wrote the initial draft. All authors subsequently critically edited the report. All authors read and approved the final report. CZ, XD, QF, HM, QZ and XW had full access to all data in the study. CZ, XD, LW and XW had final responsibility for the decision to submit for publication.

## Funding

This study was in part funded by National Natural Science Foundation of China (NSFC) (No. 61876212 and No. 61733007). The funder had no role in study design, data collection, data analysis, data interpretation, or writing of the report.

## Notes

### Competing Interest Statement

The authors have declared no competing interest.

